# Immunosequencing and epitope mapping reveal substantial preservation of the T cell immune response to Omicron generated by SARS-CoV-2 vaccines

**DOI:** 10.1101/2021.12.20.21267877

**Authors:** Damon H. May, Benjamin E. R. Rubin, Sudeb C. Dalai, Krishna Patel, Shahin Shafiani, Rebecca Elyanow, Matthew T. Noakes, Thomas M. Snyder, Harlan S. Robins

**Affiliations:** Adaptive Biotechnologies, Seattle, Washington, USA; Stanford University School of Medicine, Stanford, California, USA

## Abstract

The Omicron SARS-CoV-2 variant contains 34 mutations in the spike gene likely impacting protective efficacy from vaccines. We evaluated the potential impact of these mutations on the cellular immune response. Combining epitope mapping to SARS-CoV-2 vaccines that we have determined from past experiments along with T cell receptor (TCR) repertoire sequencing from thousands of vaccinated or naturally infected individuals, we estimate the abrogation of the cellular immune response in Omicron. Although 20% of CD4^+^ T cell epitopes are potentially affected, the loss of immunity mediated by CD4^+^ T cells is estimated to be slightly above 30% as some of the affected epitopes are relatively more immunogenic. For CD8^+^ T cells, we estimate a loss of approximately 20%. These reductions in T cell immunity are substantially larger than observed in other widely distributed variants. Combined with the expected substantial loss of neutralization from antibodies, the overall protection provided by SARS-CoV-2 vaccines could be impacted adversely. From analysis of prior variants, the efficacy of vaccines against symptomatic infection has been largely maintained and is strongly correlated with the T cell response but not as strongly with the neutralizing antibody response. We expect the remaining 70% to 80% of on-target T cells induced by SARS-CoV-2 vaccination to reduce morbidity and mortality from infection with Omicron.

## Introduction

As the SARS-CoV-2 virus continues to evolve into new strains, the amino acid sequence of the surface glycoprotein (“spike protein”) is diverging from the originally identified strain upon which all currently available COVID-19 vaccines were designed. Given the global scale of the pandemic, a vast amount of data has been collected on the immune response induced by SARS-CoV-2 vaccines, the effect of this response on each strain and the efficacy of the vaccines in each context^1–8^. Both components of the adaptive immune system, T cells and antibody-producing B cells, have been well-characterized in the setting of SARS-CoV-2 infection or vaccination^9–16^.

The SARS-CoV-2 virus appears to violate the dogma that vaccine efficacy is primarily determined by neutralization from antibodies. Despite the observation that circulating variants escape most of the neutralization induced by the vaccines, the efficacy of the available SARS-CoV-2 vaccines has remained high^2,3,6^. The SARS-CoV-2 vaccines induce a more robust T cell response than expected^17,18^, which has played a role in preventing severe (or even symptomatic) disease in vaccine recipients^19^. The T cell response has been only mildly affected by the mutations in the variants^20,21^. Surprisingly, the T cell response appears to partially offset the abrogation of neutralization from antibodies and helps control infection as well (Figure 1), with recent research demonstrating direct influence of cytotoxic T cell responses to some antigens leading to milder disease^13,15,22^.

**Figure 1.**
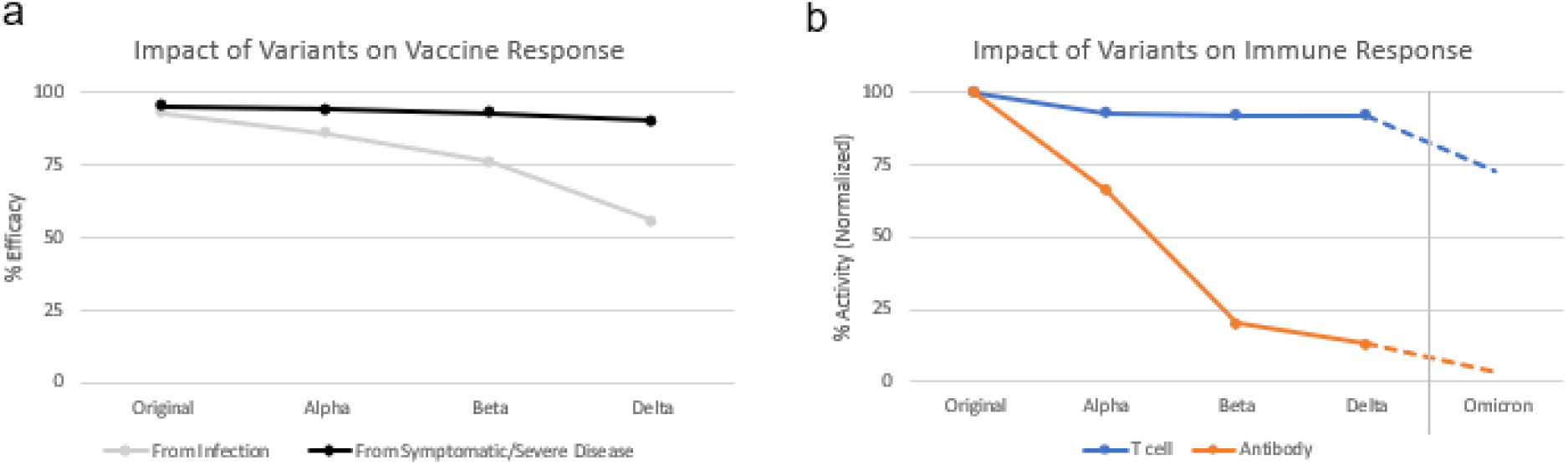
**a** Highlighting the apparent impact of variants of SARS-CoV-2 on the efficacy of vaccines, splitting out reported efficacy from infection from reported efficacy for severe or symptomatic disease. **b** Apparent impact of variants on two aspects of the immune response towards the spike protein, from T cells and from neutralizing antibody titers (normalized to the peak of response expected against the initial strain of SARS-CoV-2). Projections are made for Omicron based on early evidence and the analysis performed here. Both panels aggregate data from multiple publications and vaccines/antibody tests to give a representative view of trajectories.

Early data regarding Omicron, a new SARS-CoV-2 variant which emerged in November 2021, suggest rapid worldwide spread^23^. The Omicron variant encodes 50 protein-coding mutations in the genome, including more than 30 in the spike protein, as compared to the original SARS-CoV-2 strain used to formulate the current set of available vaccines. For comparison, the current and previous widely circulated variants contained 12 or fewer mutations in the *spike* gene. The neutralizing antibody response generated by the currently available SARS-CoV-2 vaccines was reduced against the previous Alpha, Beta, and Delta variants and several of the key efficacy-reducing mutations are also present in Omicron. Considering the continued acquisition of mutations, it is expected the antibody response will be further diminished against Omicron^24^. In contrast, prior work has shown that the vast majority of the cellular immune response induced by current vaccines remains on target against widely circulating variants including Alpha, Beta, and Delta^25^. The substantially larger set of spike mutations in the Omicron variant increases the potential to avoid the cellular immune response induced by SARS-CoV-2 vaccines. Here, we assess the fraction of both the HLA class I and class II-restricted cellular immune response impacted by the mutations in the Omicron variant.

## Results

As part of the ImmuneCODE project, we have mapped the cellular immune response to SARS-CoV-2. This mapping includes resolution of 199 HLA class I epitopes across the spike protein, with resolution to exact n-mers or short overlapping stretches, as well as resolution of the full set of HLA class II epitopes to the level of approximately 50 amino acid windows. Additionally, we have accumulated data from immunosequencing of the TCR repertoires from over 5,000 individuals following SARS-CoV-2 infection or vaccination. Combining these two sources of data allows us to determine the relative fraction of T cells induced by each antigen as well as the HLA association of each T cell clone.

Utilizing these data and a few simple assumptions, we are able to assess the impact of the mutations on cellular immune response. The primary assumption is that the mutations were not selected to avoid the cellular immune response; confidence in this assumption is supported by the diversity of HLA molecules that present the epitopes. To test this assumption, we reviewed our available data to identify if there was any significant enrichment of class I or II HLAs for prior variants and for Omicron looking across all antigen mutations. We did not observe any statistically significant enrichments from the Delta and Omicron variants within spike; however, a class II HLA association (DQA1*05:05+DQB1*03:01) connected to a nucleocapsid phosphoprotein antigen mutation (R203M in Delta and R203K/G204R in Omicron) was identified from both repertoire-focused and antigen-focused analyses. The second assumption relates to the probability that a mutation in an epitope or flanking sequence either prevents the epitope from being presented or affects binding, rendering vaccine- or infection-induced TCRs off-target. As a conservative assumption, we estimate any coding mutation spanning a presented epitope would impact all binding of TCRs to that epitope.

Given these assumptions, we determined that the mutations in the Omicron variant impact as much as 21% of the class I-restricted and 33% of the class II-restricted cellular immune response induced by SARS-CoV-2 vaccines (Figure 2, averaged projection in Figure 1B). Some mutations may not impact antigen presentation or recognition by certain TCRs, but this assessment score is an upper bound on the impact that can be inferred from available experimental data. For T cell responses targeted to non-spike regions that are induced by natural infection, the impact on the Omicron variant is miniscule (≤ 5%), because the mutations are concentrated in the *spike* gene and rare in the rest of the genome.

**Figure 2.**
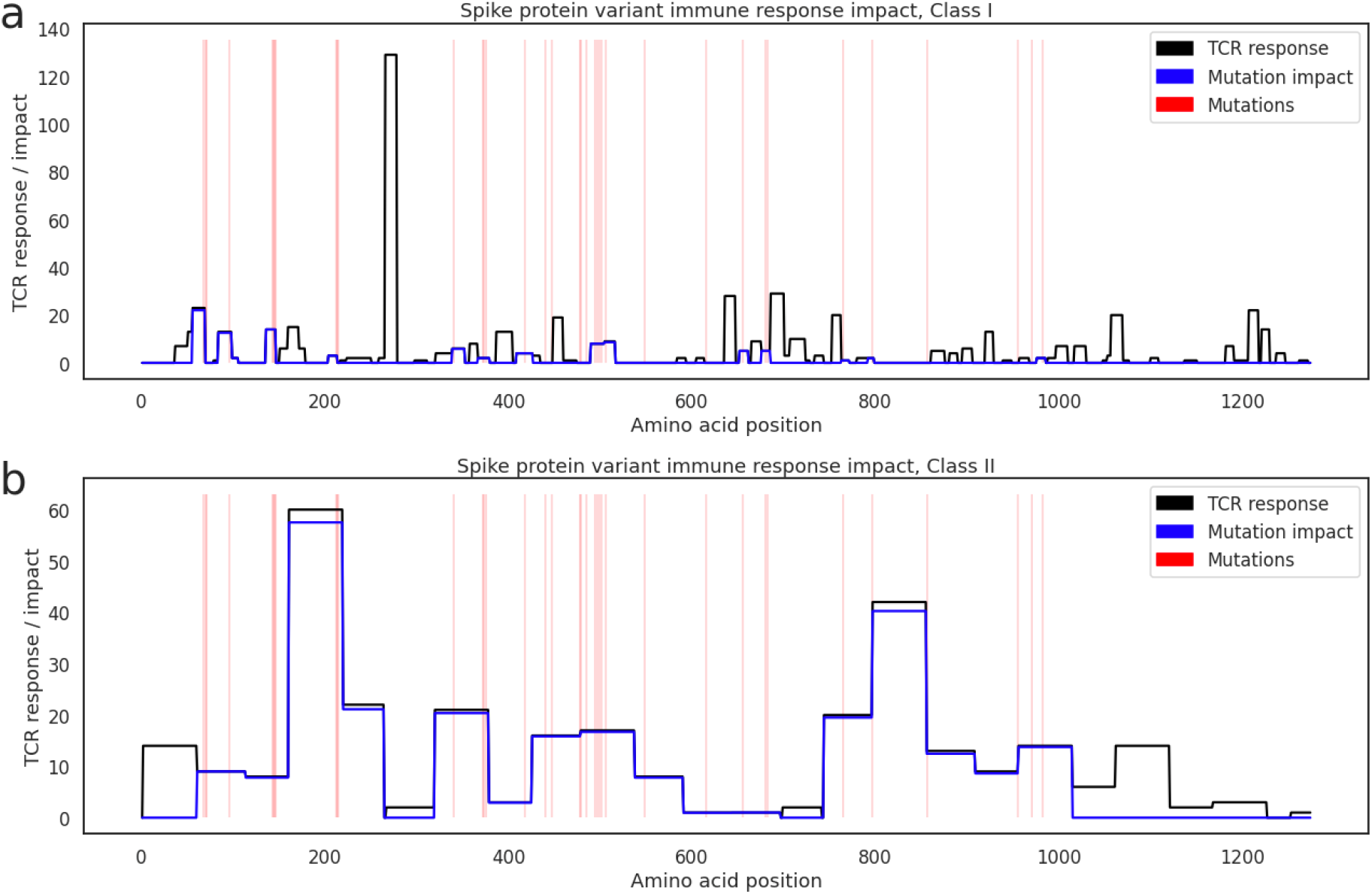
Depicting size of the Class I (**a**) and Class II (**b**) T cell response to each epitope range across the spike protein in our antigen map (black line), with positions of Omicron mutations marked in red. The blue line estimates the amount of the cellular immune response that is off-target within each epitope window when challenged with Omicron.

## Discussion

We have estimated the impact of Omicron mutations on the binding of the vaccine-induced cellular immune response, both for CD4^+^ and CD8^+^ T cells. Although there is a measurable reduction in on-target T cell memory, over two-thirds of the cellular immune response is expected to remain preserved. For individuals with prior SARS-CoV-2 infection with other variants, we expect a similar reduction in the spike-specific cellular immune memory. However, the mutations in Omicron are highly concentrated in the spike protein^26^. As the number of mutations impacting T cell epitopes in other genes of SARS-CoV-2 is comparable to prior variants with over 95% conservation, we expect almost no abrogation of the cellular immune response outside of the spike region. In particular, the CD8^+^ T cell response to natural SARS-CoV-2 infection is distributed across the full viral genome and is expected to remain well-directed and diverse against Omicron regardless of which variant was responsible for the natural infection.

Whether the T cell response to SARS-CoV-2 can prevent initial, symptomatic, mild, and/or severe infection remains an open question. The present dogma is that T cells can prevent severe and possibly mild COVID-19 illness, but likely do not prevent initial infection^27,28^. However, these considerations may be less relevant in the context of vaccine-associated prevention of SARS-CoV-2 transmission and symptomatic control. For example, it is conceivable that a mucosal T cell response could eliminate virus-infected cells along the respiratory tract sufficiently enough that an individual may experience mild or asymptomatic illness with reduced or absent risk of transmission to others and lower likelihood of testing positive for SARS-CoV-2 infection. In other words, in the setting of a robust T cell response, the distinction that antibodies can prevent while T cells resolve infection may not be as consequential.

The differences in the protective efficacy of SARS-CoV-2 vaccines predicted from evaluation of humoral versus cellular immune responses underscore the importance of T cell testing as a reliable measure of vaccine-induced immune protection, particularly in the setting of new and emerging viral variants that can evade immune surveillance. The recent development of a high-throughput, scalable assay to measure the cellular immune response to SARS-CoV-2 as demonstrated by Elyanow et al.^29^, Dalai et al.^30^, Snyder et al.^31^, and Gittelman et al.^32^ empowers such an approach, with important implications for identifying individuals and groups who may be at high risk of severe illness by new variants. As the variants continue to mutate, however, additional studies will also be needed to assess the evolving predictive value of the T-cell response.

## Methods

### Source of samples used in analysis

The data analyzed in this study were derived from samples that had been acquired and sequenced previously. The data source included a public database (GSAID) for which no IRB approval is necessary. The MIRA data were from the ImmuneRACE study^33^, Bloodworks NW (Seattle, WA), and Discovery Life Sciences (DLS) commercial vendor. The ImmuneRACE study was approved by Western Institutional Review Board (WIRB reference number 1-1281891-1, Protocol ADAP-006). Bloodworks NW donor samples had been consented and collected under the Bloodworks Research Donor Collection Protocol BT001, while the DLS samples had been collected under Protocol DLS13.

### Meta-analysis of vaccine efficacy

Vaccine efficacy estimates from literature were gathered for BNT162b2 (Pfizer)^1–3,8^, mRNA-1273 (Moderna)^3,4,8^, and Ad26.COV2.S (Janssen)^5,6^ vaccines, including the registrational studies run by these vaccine makers and real-world large scale efficacy studies organized by others. Separate measures of protection against symptomatic or severe infection, or protection from any infection, were taken based off endpoints reported across studies. Recognizing vaccine efficacy changes with time, outputs focused on the first 3 months after full vaccine administration were prioritized in aggregating data for comparison to variants.

### Meta-analysis of antibody activity against variants

Neutralizing antibody and other antibody readouts were gathered from multiple published studies, including studies that included DFT (IC50) measures to allow some comparison across vaccines, primarily from Yu et al.^34^ and supported by Sadoff et al.^6^, Jackson et al.^35^, Folegatti et al.^36^ and Walsh et al.^37^. The fold change of neutralizing antibody response to these variants compared to the original strain was gathered for each vaccine from other publications: BNT162b2 (Pfizer) from Anichini et al.^9^, Edara et al.^10^, with support from Tada et al.^12^ and Garcia-Beltran et al.^38^; for mRNA-1273 (Moderna) from Wu et al.^11^ Edara et al.^10^ with support from Wang et al.^39^ and Garcia-Beltran et al.^38^; for Ad26.COV2.S (Janssen), Tada et al.^12^ and Alter et al.^13^ with support from Naranbhai et al.^40^. While there was some missing data for given vaccines and variants, the meta-analysis revealed consistent trends in drop-offs for variants when averaging the fold-change results.

### Mutation identification for Omicron

A total of 172 published sequences for individuals infected with Omicron in GISAID (available on Nov 30, 2021; https://www.gisaid.org/hcov19-variants/) were aligned and mutations seen in over 75% of the individuals retained as the reference sequence for Omicron. This preliminary definition has remained consistent with the now >1,000 sequences available for analysis.

### Analysis of T cell activity against variants including Omicron

To estimate the proportion of the cellular immune response impacted by the Omicron mutations, we overlapped the Omicron spike mutation list with each antigen using a convalescent COVID donor-based MIRA/antigen mapping dataset in ImmuneCODE and its associated T cell count as a measure of immunogenicity. For individually-addressed antigens in CD8^+^ T cells, a peptide with any mutation was considered to remove 100% of the response for that antigen. For multiply-addressed antigens in CD4^+^ or CD8^+^ T cells, we calculated the proportion of the peptides within each set of antigens presented together that contain one or more Omicron mutations and used that fraction to calculate an impact score. Across all antigen positions, we then estimated the total cellular immune impact over that region as the number of T cells responding multiplied by the proportion of the cellular immune response impacted, shown as the blue lines in Figure 2. The final estimates of CD4^+^ and CD8^+^ T cell reduced efficacy reflect the impacted count of T cells over the original count of T cells for all spike antigens.

## Data Availability

As part of the ImmuneCODE data resource, the COVID-19 MIRA data and COVID-19 study immunosequencing data are freely available for analysis and download from the Adaptive Biotechnologies immuneACCESS site under the immuneACCESS Terms of Use at https://clients.adaptivebiotech.com/pub/covid-2020. This resource is being updated with the additional vaccine repertoire and other data included in this analysis.

## Data Availability

As part of the ImmuneCODE data resource^41^, the COVID-19 MIRA data and COVID-19 study immunosequencing data are freely available for analysis and download from the Adaptive Biotechnologies immuneACCESS site under the immuneACCESS Terms of Use at https://clients.adaptivebiotech.com/pub/covid-2020. This resource is being updated with the additional vaccine repertoire and other data included in this analysis. ImmuneCODE is for research use only and is not for use in diagnostic procedures.

## Acknowledgments

The ImmuneCODE data resource that underlies this analysis paper is the result of collaboration among many individuals and organizations working together to advance global understanding of SARS-CoV-2 and COVID-19. We are grateful for the support and participation of all our partners. We are especially grateful for the generosity of the participants who donated blood for this and other studies.

We would like to thank Rachel Gittelman and Mark Klinger for helpful discussions and review of some of these findings. We would like to thank Kristin MacIntosh for editorial support.

